# The impact of neighborhood factors on exercise capacity in children with hypertrophic cardiomyopathy

**DOI:** 10.1101/2024.10.22.24315501

**Authors:** Imran R. Masood, Lei Wang, Helen M. Stanley, Jonathan J. Edwards, Humera Ahmed, Kimberly Y. Lin, Carol A. Wittlieb-Weber, Matthew J. O’Connor, Joseph W. Rossano, Shannon O’Malley, Stephen Paridon, Vicky W. Tam, Jonathan B. Edelson

## Abstract

**Background:** Restricting certain patients with hypertrophic cardiomyopathy (HCM) from exercise likely has negative cardiovascular effects and may not reduce the risk of sudden cardiac death. Promoting exercise in children with HCM is complex and requires knowledge of the environmental factors that impact exercise capacity in children with HCM.

**Methods:** This retrospective, cross-sectional analysis includes children with HCM who underwent exercise stress testing (EST) at a single, children’s tertiary-care center between 2000 and 2023. Addresses from contemporaneous EST were accessed and geocoded to census tracts. The Child Opportunity Index (COI) was the primary exposure of interest. Granular neighborhood measures including walkability index, Rural-Urban Commuting Area Codes (RUCA), Index of Concentration at the Extremes and Uniform Crime Reporting rates were measured. The primary outcome measure was peak oxygen consumption (VO_2_). Linear regression and multivariable analyses were performed.

**Results:** A total of 155 patients were identified who met inclusion criteria, 23% (n=35) of whom were female. Mean age at time of EST was 15.8 ±3.1 years. More than half of included patients were from high or very high COI (30%, n=46, and 35%, n=54, respectively). Most patients lived in urban environments (RUCA score 1 or 2, 96.7%, n=150). The mean peak VO_2_ was 2159 ±906 milliliters/min and adjusted peak VO_2_ 35.5 ±9.3 mL/kg/min. A multivariate model adjusting for disease severity, age at diagnosis of HCM, race and accounting for collinearity, showed that low COI, higher levels of urbanization and lower concentration of neighborhood wealth were independently associated with lower peak VO_2_.

**Discussion:** Our study identified previously unrecognized environmental determinants of exercise capacity in children with HCM, with lower COI, increased urbanization, and lower neighborhood wealth independently associating with lower exercise performance. Programs designed to increase physical activity levels and exercise performance in children with HCM should account for neighborhood and economic factors.

## Introduction

Traditionally, pediatric patients with hypertrophic cardiomyopathy (HCM) have been restricted from physical activity due to the perceived risk of sudden cardiac death with exertion^1^. However, contemporary data have demonstrated that physical activity may be safe for certain patients with HCM, and that exercise restriction is associated with negative cardiovascular and mental health consequences^2,3^. Of additional concern is that the effect of activity restriction appears cumulative, with data demonstrating a decline in exercise capacity with increasing age^4^.

While these data highlight the importance of developing interventions that promote safe physical activity in children with HCM, the design and implementation of physical activity-related interventions are complex. For interventions to be effective, there must be an understanding of determinants of exercise capacity. Adapting Bronfenbrenner’s ecological systems theory^5^, we can understand exercise capacity in children with HCM as being influenced by a diverse range of factors including individual factors (age, illness severity^4,6^, medical comorbidities), parent and healthcare provider attitudes^7,8^, and the environment in which the child lives.

However, the relative contributions of these factors are not well defined, and this is a major impediment to the design of interventions aimed at increasing levels of physical activity. To help address the knowledge gap related to the determinants of exercise, we focused on how environmental factors may influence exercise capacity in children with HCM using a retrospective cohort design.

A useful framework for describing a child’s environment is to divide the area they live into two interrelated, but separate, spheres: the built and the natural. The built environment is characterized by the urban planning of a neighborhood (sidewalks, public transportation), recreational access (gyms, playgrounds), and greenspaces (parks, etc.),^9^ while the natural environment includes naturally occurring elements such as vegetation, bodies of water and climate. Prior investigations have shown that features of the built environment, including traffic density, sidewalk access, and the presence of speed bumps and pedestrian walk signals, impact physical activity levels in healthy children^10–12^, and in neighborhoods with high crime rates, children are less likely to be active^13^. Furthermore, in socioeconomically advantaged communities, there are different opportunities to engage in community or organized sports, or access recreational facilities^11^. The relationships of these factors to exercise capacity in children with HCM has not been evaluated and is likely to be different given the related impact of differential access to healthcare resources for these children. Based on this conceptual model (figure 1), we hypothesized that the natural and built environments in which children with HCM live will represent a critical and underrecognized role in determining their exercise capacity and ultimately their levels of physical activity.

**Figure 1.**
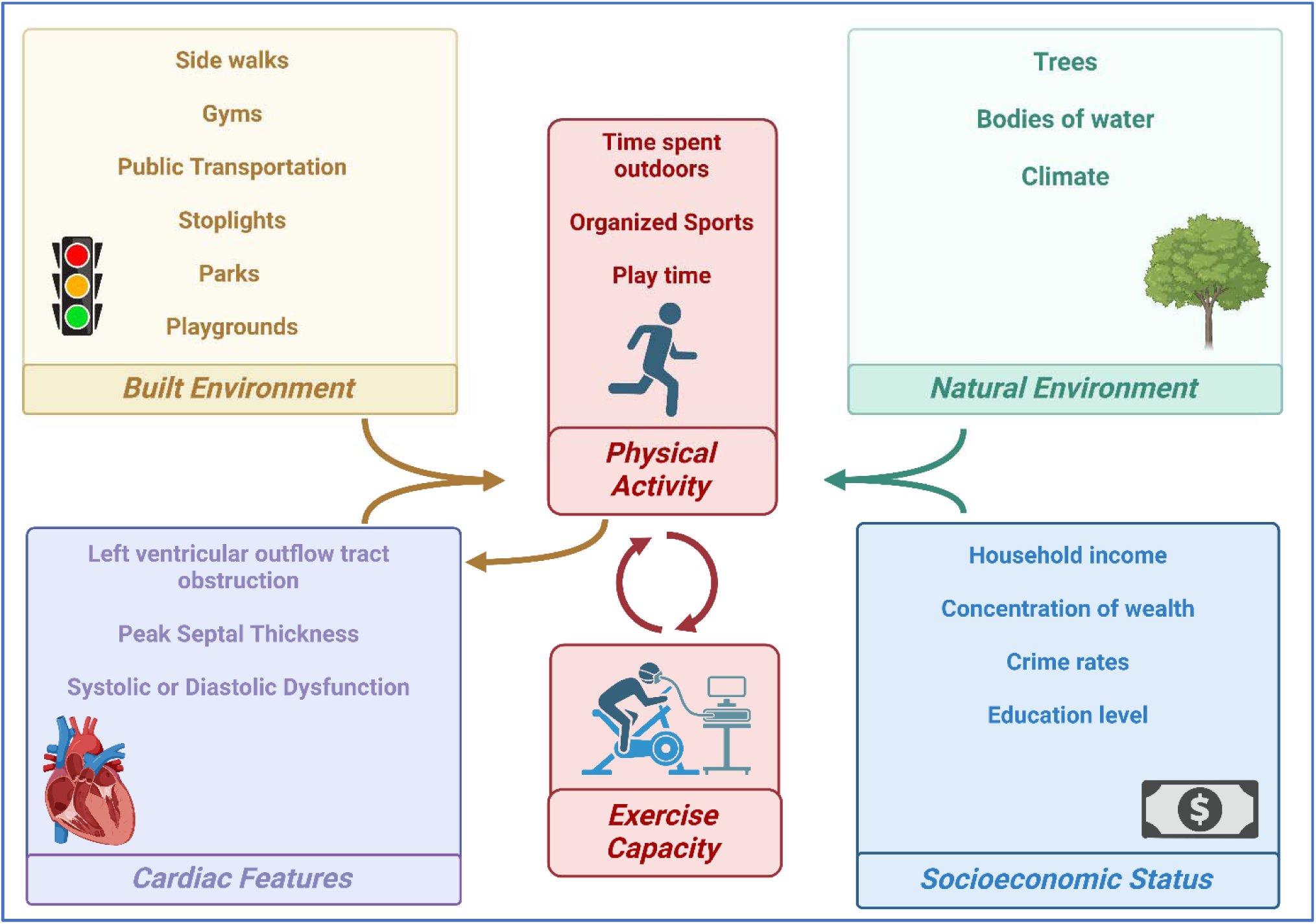
Conceptual model of multiple determinants of exercise capacity in children with hypertrophic cardiomyopathy.

## Methods

### Study design

This was a retrospective, cross-sectional study performed at a tertiary care children’s hospital of patients <21 years old with a diagnosis of HCM who underwent cardiopulmonary exercise testing (EST) between the years 2000 and 2023. It is customary practice at our institution that adolescents with HCM undergo annual EST at physician discretion. Potential subjects were identified by querying a longitudinal internal database that includes all exercise tests performed at our institution and identifying those patients with a diagnosis code for HCM. Subjects’ charts were evaluated to confirm a correct diagnosis of HCM and to abstract additional clinical and demographic data. Subjects designated as either genotype-positive/phenotype-negative or ‘possible HCM’ were excluded from the study cohort. Subject addresses from the time of the contemporaneous EST were identified and abstracted by utilizing the home address included on billing forms generated at the time of EST. Patient home addresses were then geocoded and spatially linked to census block groups to serve as a proxy for patient neighborhood. This allows us to describe and evaluate the environmental exposures listed below. Demographic data including the subject’s age at the time of exercise stress testing, sex, and race were also collected. This study was approved by the Institutional Review Board at the Children’s Hospital of Philadelphia. Written informed consent was waived given the retrospective nature of this study.

### Covariates

Electronic medical record abstraction was used to record each patient’s genetic diagnosis if present, underlying mutation, cardiac medications, and echocardiography data. These covariates were selected given their relationship to disease severity. Peak septal thickness was measured using echocardiography M-mode. Z-scores for echocardiographic parameters were normalized to body surface area and were considered normal if they were within two standard deviations of the population mean. The echocardiogram performed most closely in time to the EST was included in the analysis. Stress echocardiography data was not included in this study.

### Exposures of interest

The primary exposure of interest was the Child Opportunity index 3.0 (COI)^14^. The COI is a census tract level measure of neighborhood resource quality and conditions leading to healthy child development and economic opportunity, broadly reflecting both the natural and built environments. There are three main domains within COI: (1) education, (2) health and environment and (3) social and economic, comprising 44 indicators. Data from the years 2012 to 2021 were included. Each nationally normed COI category is ranked numerically from 1-100, with 100 representing a neighborhood with the highest opportunities for children. COI domains are also categorically ranked as very low, low, moderate, high or very high. After evaluating the distribution of this exposure in our sample, we grouped COI into two categories: low (including very low and low), or high (including moderate, high and very high). COI data from 2012-2022 was available, therefore data with the closest date to the patient’s EST was chosen for analysis. The COI was chosen based on its comprehensive nature, and its organization into three separate domains. We anticipated identifying different associations of the domains of COI (education, health/environment, and social economic domains) with exercise capacity that could guide the next steps in this investigative pathway.

In selecting more granular aspects of the built environment, we chose to measure the Walkability index (WI)^15^ (data from year 2019), Rural-Urban Commuting Area Codes (RUCA)^16^ (data from year 2010), Index of Concentration at the Extremes (ICE)^17^ (data from year 2020), Normalized Difference Vegetation Index (NDVI)^18^ (data from year 2022) and Uniform Crime Reporting (UCR) rates^19^ (data from year 2014). These environmental exposures were selected given the key components of our conceptual model. We hypothesized that children with the ability to spend time outdoors playing, walking, or running would have better exercise capacity due to having higher physical activity levels, and therefore that neighborhood traffic, crime rates, and access to green space, sidewalks, or walking paths would impact physical activity. Furthermore, we theorized that children from wealthier neighborhoods would have higher physical activity levels by having better access to recreational facilities and being more likely to join organized sports^11^. These indicators offer value in addition to the COI, given their more granular measurement of factors that purportedly impact physical activity.

The WI is a tool created by the United States Environmental Protection Agency (EPA) that assesses how conducive an area is to walking. For each block group, the WI is ranked on a scale of 1 to 20, 1 to 5.57 being the least walkable, 5.76 to 10.5 being below average walkable, 10.51 to 15.25 being above average walkable, and 15.26 to 20 being the most walkable. RUCA is a tract-level measure of urbanization, population density, and daily commuting, which is rated on a scale of 1 to 10, where 1 represents the most urban environment, and 10 the most rural. Developed by the U.S. Department of Agriculture (USDA), RUCA codes help in understanding the extent of rurality and urbanity for census tracts. NDVI is a metric used to assess vegetation and vegetation health by comparing the difference between near-infrared and red-light reflectance. We used high-resolution aerial imagery from The National Agricultural Imagery Program (NAIP) from 2022 to calculate the average NDVI value for each tract. NDVI ranges from −1 to +1, with −1 to 0 indicating cloud cover, urbanized area, or water, values near 0 indicating bare soil, and higher values indicating dense, green vegetation. The ICE measures the degree of structural racism by quantifying extremes of deprived or privileged social groups by accounting for income inequality and racial/ethnic segregation at the census tract level. ICE is valued continuously from –1 to +1, with –1 signifying a concentration of societal deprivation, and +1 signifying a concentration of societal advantage. ICE is calculated using the difference between the number of advantaged people and disadvantaged people, divided by the total number living in that community. Total crime rates were obtained from the uniform crime reporting program, provided by the Federal Bureau of Investigation (FBI). Crime data was reported at the county level, and contained various rate subtypes including murder, rape, robbery, aggravated assault, total property crimes, burglary, larceny, and motor vehicle theft. The aforementioned neighborhood exposures were selected given their established relationships with physical activity levels in healthy children^13,20,21^.

### Outcome measure

The primary outcome measure was exercise capacity, defined as the peak oxygen consumption (VO_2_) achieved during EST. Peak VO_2_ is a marker of cardiovascular fitness and is a potentially useful surrogate marker for physical activity levels. Furthermore, peak VO_2_ is often utilized as an endpoint for exercise interventions and plays an important investigational role in pediatric cardiomyopathy. Using a widely available and generalizable outcome measure naturally lends itself to the development of future interventional studies. Exercise stress testing data was obtained by electronic medical record abstraction. Exercise testing was performed using a standard watt-ramp protocol on cycle ergometry or treadmill testing using protocols designed at the Children’s Hospital of Philadelphia. At our institution, cycle ergometry is the default unless the patient is too short to use the ergometer correctly. All ESTs were performed by an exercise physiologist and supervised by a pediatric cardiologist. Patients with HCM are typically referred for exercise testing for routine surveillance, to assess functional capacity, to measure exercise-induced arrhythmias, or for pre-sports participation risk assessment. Metabolic data was obtained using a Vmax Encore metabolic cart (Carefusion, Yorba Linda, California) or a Vyntus metabolic cart (Vyaire ^TM^, Mettawa, Illinois), and normative data was determined using the Cooper reference values^22^. Exercise testing data included the peak VO_2_, VO_2_ at anerobic threshold, work rate (watts), and the heart rate response. VO_2_ was adjusted for weight using ideal body weight and was expressed as milliliters/kilogram/minute (mL/kg/min). Heart rate reserve was defined as the difference between the expected peak heart rate and the observed peak heart rate. If multiple ESTs were present for a single subject, the most recent EST was used for the statistical analysis.

### Statistical Analysis

The statistical analysis performed in this study involved presenting continuous data as arithmetic mean and standard deviation (SD), while categorical data were presented as frequency. The main aim was to examine the associations between neighborhood exposures and exercise capacity, with linear regression utilized as the primary analytical method. In the initial analysis, a simple linear regression model was employed to measure the impact of neighborhood exposures and cardiac features on peak VO_2_.

Subsequently, an additional analysis was conducted, wherein VO_2_ was regressed on the overall COI along with other non-COI predictors including the walkability index, RUCA, total violent crime rate, total property crime rate, NDVI, index of extremes, BMI, age at diagnosis of HCM, patient race, septal thickness, and left ventricular outflow tract gradient.

A statistically significant finding was defined a priori as one with a p-value < 0.05. This threshold was used to determine the significance of predictor variables in both the initial and adjusted regression models, helping to identify factors significantly associated with exercise capacity after accounting for potential confounders. Subjects in which neighborhood exposures could not be fully obtained or in those with incomplete co-variate data were removed from the sample.

## Results

### Demographic and clinical data

After querying diagnosis codes and removing those without a diagnosis of HCM, and one patient with a Canadian address, there were 170 patients who met inclusion criteria. Complete neighborhood data was available for 155. The mean age at the time of exercise testing was 15.8 ± 3.1 years, with 23% (n=35) female patients. Mean body mass index was elevated at 32.7 ± 25.0. The most common race was white (70%, n=109). The mean age at diagnosis of HCM was 10.8 ± 5.6 years. A full description of demographic and clinical data is shown in table 1.

**Table 1.** Demographics and Clinical Characteristics.

| **Demographics** | **n = 155** |
| --- | --- |
| Age in years at EST, mean (SD) | 15.81 (3.1) |
| Age in years at diagnosis of HCM, mean (SD) | 10.77 (5.6) |
| Sex, n (%) |  |
| Male | 120 (77) |
| Female | 35 (23) |
| Ethnicity, n (%) |  |
| Not Hispanic or Latino | 149 (96) |
| Hispanic or Latino | 5 (3.2) |
| Unknown | 1 (0.6) |
| Race, n (%) |  |
| White | 109 (70) |
| Black or African American | 37 (24) |
| Asian | 5 (3.2) |
| Unknown / Not Reported | 4 (2.6) |
| **Clinical Characteristics** |  |
| LV ejection fraction by echo measured as percent, mean (SD) | 66.92 (8.0) |
| Peak LVOT gradient by echocardiogram in mmHg, mean (SD) | 26.50 (50.4) |
| Mean LVOT gradient by echocardiogram in mmHg, mean (SD) $\alpha$ | 3.14 (8.3) |
| Maximum interventricular septal diameter in mm, mean (SD) | 1.72 (0.8) |
| Maximum interventricular septal diameter Z-score, mean (SD) | 5.32 (5.4) |
| Cardiac Medications, n (%) |  |
| Beta blocker | 66 (43) |
| Beta blocker and CCB | 2 (2.6) |
| Beta blocker, CCB and Disopyramide | 3 (3.9) |
| CCB | 3 (3.9) |
| CCB and Disopyramide | 1 (1.3) |
| Disopyramide | 1 (1.3) |
| Unknown | 79 (51) |
| Genetic Mutation or Syndrome present, n (%) |  |
| Yes | 77 (50) |
| No | 76 (50) |

EST = exercise stress test. HCM = hypertrophic cardiomyopathy. CCB = calcium channel blocker. LV = left ventricle. LVOT = left ventricular outflow tract.
$\alpha$ Data available for 88 patients

### Covariate data

By echocardiography, the mean peak septal thickness was 1.7 ± 0.8 cm (mean Z-score +5.32 ± 5.4), and the mean peak instantaneous left ventricular outflow tract Doppler gradient was 26.5 ± 50 mmHg. Beta-blockers were used in 43% (n=66) of patients. The mean ejection fraction was 66.9% ± 8.0.

### Neighborhood data

There were 7 different states (Pennsylvania, New Jersey, Delaware, Maryland, Virginia, Michigan and Florida) and 121 distinct tracts seen, of which 9.0% (n = 14) were Philadelphia tracts. More than half of the included patients were from high or very high overall COI (30%, n = 46, and 35%, n = 54, respectively), suggesting most patients lived in neighborhoods with high opportunity. The distribution of COI categories is shown in figure 2. The mean overall COI score was 65.2 ± 26.7 and the mean overall COI Z-score was 0.50 ± 0.7. Most patients in this sample lived in an urban environment, as represented by the RUCA score (96.7%, n = 150 for RUCA score of 1 or 2). The majority of patients had either below average or above average walkability (38%, n = 59 and 34%, n = 52, respectively). Neighborhood exposure data is shown in table 2.

**Figure 2.**
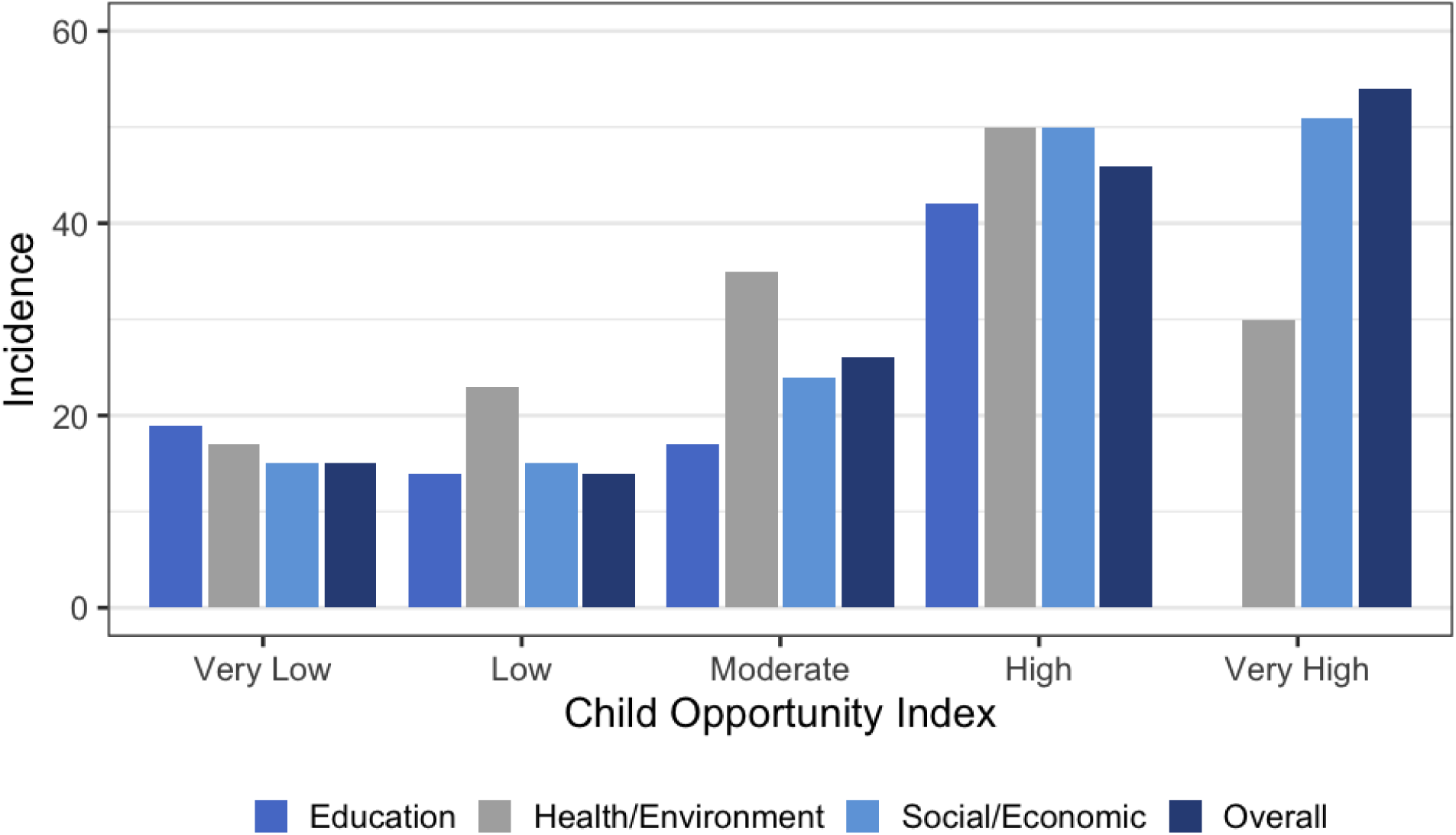
Distribution of Child Opportunity Index.

**Table 2.**
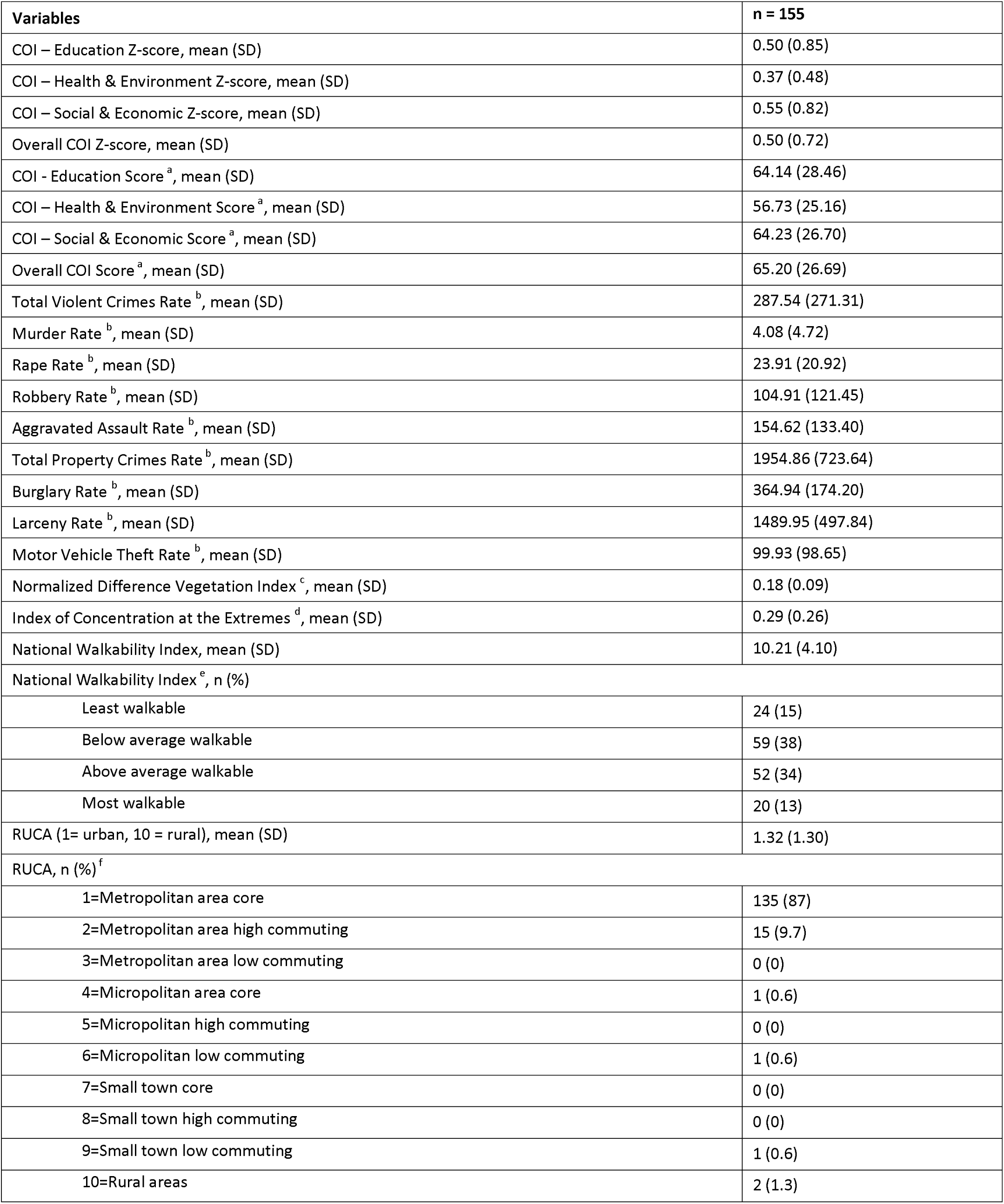
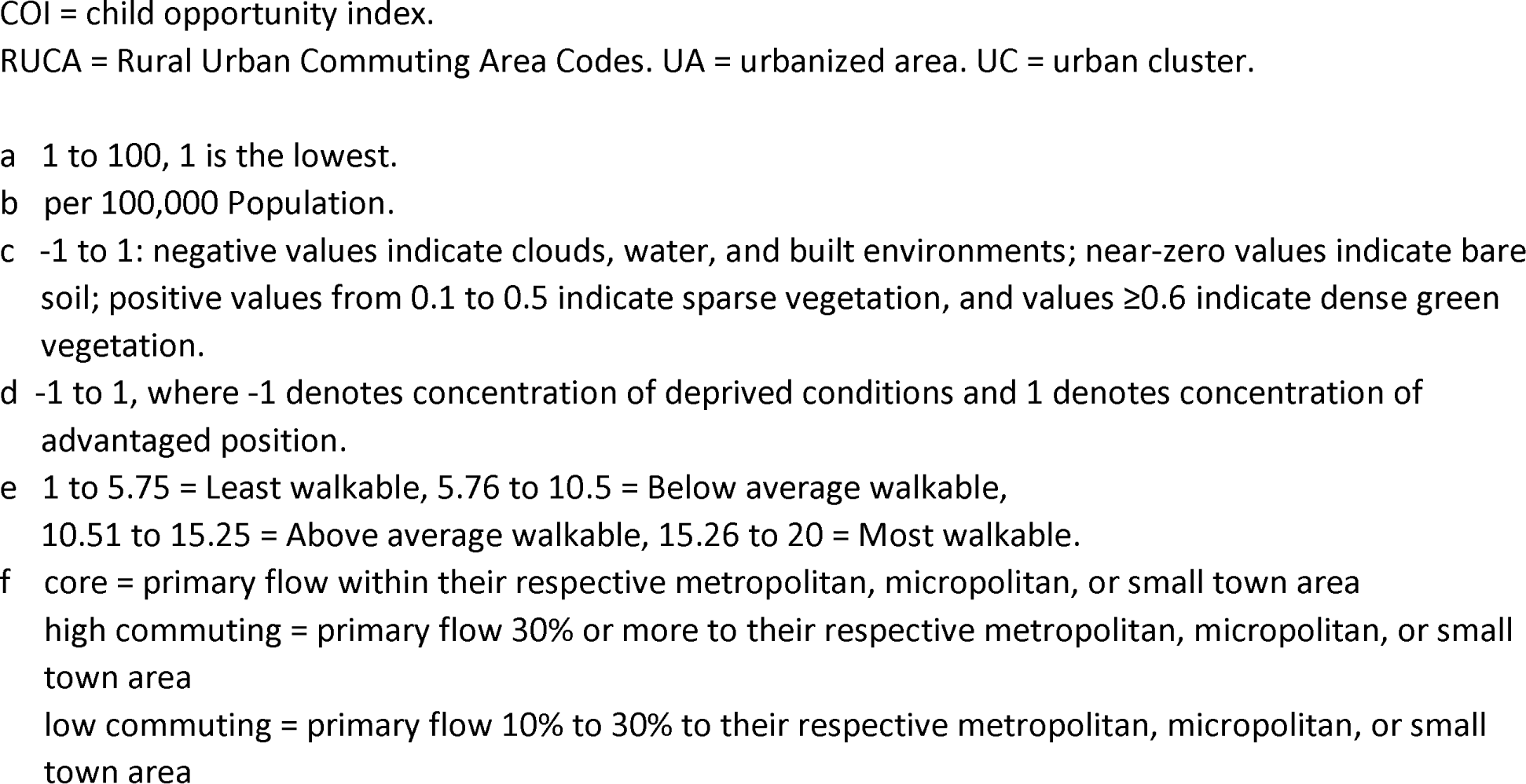
Neighborhood Data.

| **Variables** | **n = 155** |
| --- | --- |
| COI – Education Z-score, mean (SD) | 0.50 (0.85) |
| COI – Health & Environment Z-score, mean (SD) | 0.37 (0.48) |
| COI – Social & Economic Z-score, mean (SD) | 0.55 (0.82) |
| Overall COI Z-score, mean (SD) | 0.50 (0.72) |
| COI - Education Score <sup>a</sup>, mean (SD) | 64.14 (28.46) |
| COI – Health & Environment Score <sup>a</sup>, mean (SD) | 56.73 (25.16) |
| COI – Social & Economic Score <sup>a</sup>, mean (SD) | 64.23 (26.70) |
| Overall COI Score <sup>a</sup>, mean (SD) | 65.20 (26.69) |
| Total Violent Crimes Rate <sup>b</sup>, mean (SD) | 287.54 (271.31) |
| Murder Rate <sup>b</sup>, mean (SD) | 4.08 (4.72) |
| Rape Rate <sup>b</sup>, mean (SD) | 23.91 (20.92) |
| Robbery Rate <sup>b</sup>, mean (SD) | 104.91 (121.45) |
| Aggravated Assault Rate <sup>b</sup>, mean (SD) | 154.62 (133.40) |
| Total Property Crimes Rate <sup>b</sup>, mean (SD) | 1954.86 (723.64) |
| Burglary Rate <sup>b</sup>, mean (SD) | 364.94 (174.20) |
| Larceny Rate <sup>b</sup>, mean (SD) | 1489.95 (497.84) |
| Motor Vehicle Theft Rate <sup>b</sup>, mean (SD) | 99.93 (98.65) |
| Normalized Difference Vegetation Index <sup>c</sup>, mean (SD) | 0.18 (0.09) |
| Index of Concentration at the Extremes <sup>d</sup>, mean (SD) | 0.29 (0.26) |
| National Walkability Index, mean (SD) | 10.21 (4.10) |
| National Walkability Index <sup>e</sup>, n (%) |  |
| Least walkable | 24 (15) |
| Below average walkable | 59 (38) |
| Above average walkable | 52 (34) |
| Most walkable | 20 (13) |
| RUCA (1= urban, 10 = rural), mean (SD) | 1.32 (1.30) |
| RUCA, n (%) <sup>f</sup> |  |
| 1=Metropolitan area core | 135 (87) |
| 2=Metropolitan area high commuting | 15 (9.7) |
| 3=Metropolitan area low commuting | 0 (0) |
| 4=Micropolitan area core | 1 (0.6) |
| 5=Micropolitan high commuting | 0 (0) |
| 6=Micropolitan low commuting | 1 (0.6) |
| 7=Small town core | 0 (0) |
| 8=Small town high commuting | 0 (0) |
| 9=Small town low commuting | 1 (0.6) |
| 10=Rural areas | 2 (1.3) |

COI = child opportunity index.
RUCA = Rural Urban Commuting Area Codes. UA = urbanized area. UC = urban cluster.
- a 1 to 100, 1 is the lowest. - b per 100,000 Population. - c -1 to 1: negative values indicate clouds, water, and built environments; near-zero values indicate bare soil; positive values from 0.1 to 0.5 indicate sparse vegetation, and values $\geq 0.6$ indicate dense green vegetation. - d -1 to 1, where -1 denotes concentration of deprived conditions and 1 denotes concentration of advantaged position. - e 1 to 5.75 = Least walkable, 5.76 to 10.5 = Below average walkable, 10.51 to 15.25 = Above average walkable, 15.26 to 20 = Most walkable. - f core = primary flow within their respective metropolitan, micropolitan, or small town area high commuting = primary flow 30% or more to their respective metropolitan, micropolitan, or small town area low commuting = primary flow 10% to 30% to their respective metropolitan, micropolitan, or small town area

### Outcome measure and exercise testing

Cycle ergometry testing was used in 136 patients, and treadmill for 19 patients. The mean peak VO_2_ was 2159± 906 milliliters/min, adjusted peak VO_2_ was 35.5 ± 9.3 mL/kg/min and VO_2_ percent-predicted of 80.45 ± 20.5. There was a mean work rate of 155.8 ± 63 watts. The mean heart rate reserve was 118 ± 43. Data from exercise testing results are shown in table 3.

**Table 3.** Exercise Testing Results.

| **Description, mean (SD)** | **n = 155** |
| --- | --- |
| Height in cm | 165.89 (14.35) |
| Weight in kg | 71.07 (25.32) |
| BMI | 32.74 (25.04) |
| Peak VO<sub>2</sub> in mL | 2159.66 (906.05) |
| Peak VO<sub>2</sub> adjusted in mL/kg/min | 35.52 (9.32) |
| Peak VO<sub>2</sub> percent of predicted | 80.45 (20.48) |
| VO<sub>2</sub> at Anaerobic threshold | 21.61 (5.78) |
| VO<sub>2</sub> at Anaerobic threshold percent of predicted | 60.56 (9.30) |
| Work rate in watts | 155.75 (62.58) |
| HR rest | 68.65 (13.56) |
| HR peak | 174.11 (24.68) |
| HR reserve | 117.76 (43.04) |
| O<sub>2</sub> pulse at peak exercise | 7.15 (14.34) |
| O<sub>2</sub> pulse at peak exercise, percent of predicted $\alpha$ | 37.81 (158.58) |

**Table 4.**
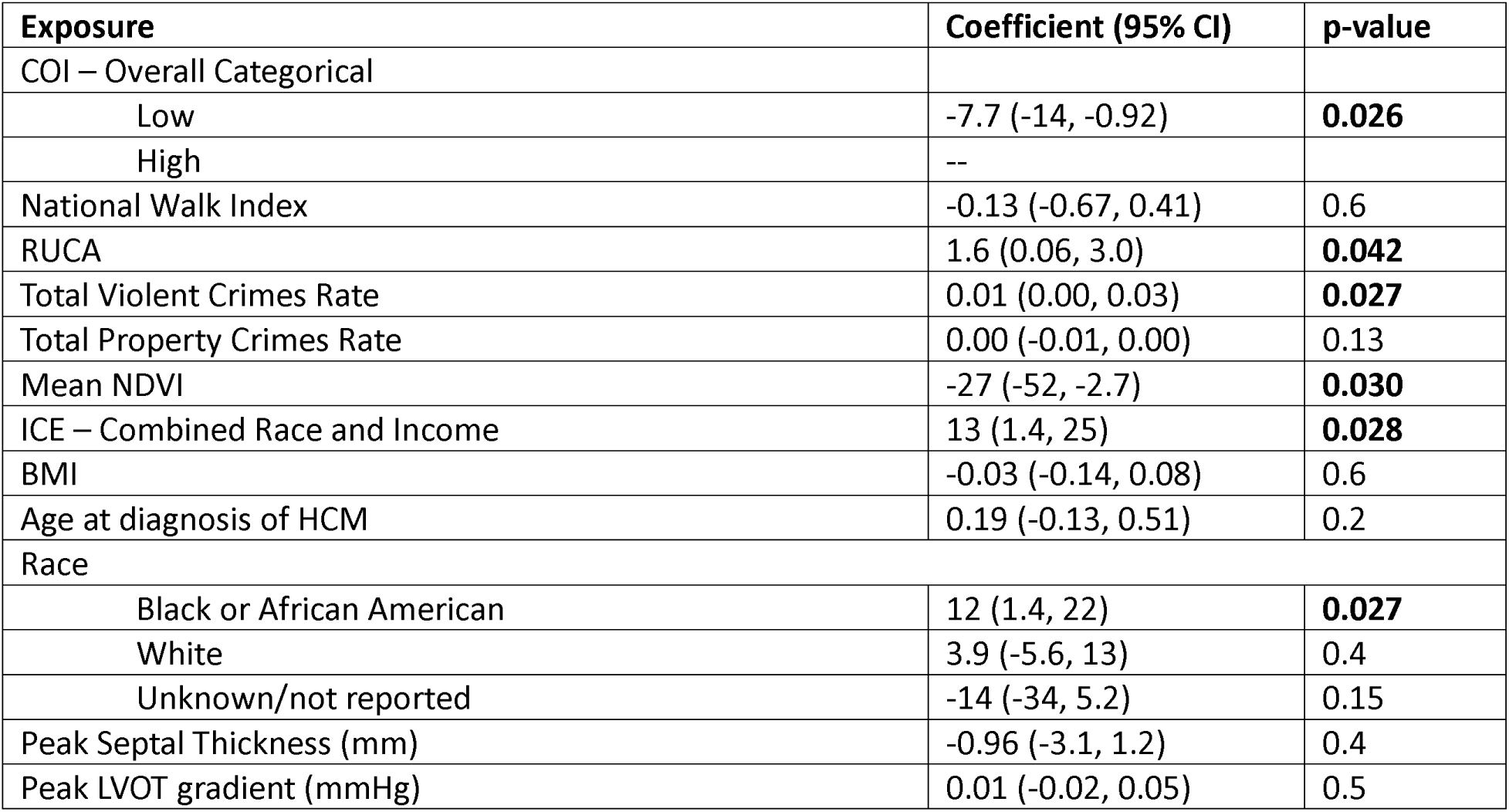
Multivariable Model to Predict Adjusted Peak VO_2_ (mL/kg/min)

| **Exposure** | **Coefficient (95% CI)** | **p-value** |
| --- | --- | --- |
| COI – Overall Categorical |  |  |
| Low | -7.7 (-14, -0.92) | **0.026** |
| High | -- |  |
| National Walk Index | -0.13 (-0.67, 0.41) | 0.6 |
| RUCA | 1.6 (0.06, 3.0) | **0.042** |
| Total Violent Crimes Rate | 0.01 (0.00, 0.03) | **0.027** |
| Total Property Crimes Rate | 0.00 (-0.01, 0.00) | 0.13 |
| Mean NDVI | -27 (-52, -2.7) | **0.030** |
| ICE – Combined Race and Income | 13 (1.4, 25) | **0.028** |
| BMI | -0.03 (-0.14, 0.08) | 0.6 |
| Age at diagnosis of HCM | 0.19 (-0.13, 0.51) | 0.2 |
| Race |  |  |
| Black or African American | 12 (1.4, 22) | **0.027** |
| White | 3.9 (-5.6, 13) | 0.4 |
| Unknown/not reported | -14 (-34, 5.2) | 0.15 |
| Peak Septal Thickness (mm) | -0.96 (-3.1, 1.2) | 0.4 |
| Peak LVOT gradient (mmHg) | 0.01 (-0.02, 0.05) | 0.5 |

### Association of neighborhood exposures with exercise capacity

In our multivariate model adjusting for disease severity, age at diagnosis of HCM, race and accounting for collinearity, we found that low COI, higher levels of urbanization (low score on RUCA) and lower concentration of neighborhood wealth were all independently associated with a lower peak VO_2_. In contrast to the directionality of many other associations, we found a positive association between county-level crime and peak VO_2_. Additionally, there was a positive correlation between Black race and peak VO_2_, independent of other neighborhood factors and disease severity. Markers of illness severity, including LVOT peak gradient and peak septal thickness were not significant predictors of peak VO_2_ in the multivariate model.

## Discussion

To our knowledge, this is the first study to analyze the impact of neighborhood factors on exercise capacity in children with HCM and provides a novel methodology for understanding previously unrecognized determinants of exercise capacity in this population. Several of these findings deserve additional discussion including: 1) children with HCM from neighborhoods with low COI have associated lower exercise capacity, 2) higher levels of urbanization correlate with worse exercise capacity, and 3) children living in less wealthy areas have associated lower exercise capacity. These findings have critical implications for developing interventions to promote exercise that are successful, accessible, and equitable.

Our finding that children with HCM who live in low COI neighborhoods had worse exercise capacity is important because it illustrates the utility of this in metric in capturing the varied and diverse contributors that define neighborhood resources and their collective impact on a modifiable factor of outcomes. Prior studies reveal that neighborhoods with higher COI offer better access to parks, playgrounds and sports facilities, which promote physical activity and sports participation^23,24^. Furthermore, children who attend schools in more affluent neighborhoods are more likely to be exposed to school-based programs that promote exercise and physical activity^25^. We also found that children living in more urban neighborhoods had worse exercise capacity, regardless of measures of socioeconomic status. It has been previously established that healthy children living in urban environments tend to have less access to parks, playgrounds and greenspace^26^ and lower rates of physical activity^27–29^. Furthermore, urban neighborhoods may be more dangerous with higher traffic density and less access to safe walkways^13,30,31^.

While the design of this study does not allow us to identify the mechanism behind the observed observation of these environmental factors and exercise capacity in children with HCM^11^, these findings should be taken into account when creating exercise prescriptions or developing programs to promote physical activity. One promising method for the delivery of physical activity-related interventions includes the use of virtual platforms. Prior work has shown that virtual exercise interventions can be beneficial for children with a variety of heart diseases, including improvement in markers of physical health (e.g. 6-minute walk distance and physical strength), and may overcome some of the inherent inequity imposed by the environment in which the child lives and their family’s resources ^32–34^. Additionally, the data from this study supports the critical need for public health interventions and favorable public policies aimed at making urban areas more child-friendly, particularly for those in low COI environments. Prior studies have shown that poorly-maintained spaces in urban neighborhoods are associated with high crime rates^35–38^ and fear of crime^39,40^, which deters outdoor physical activity. To modify the negative effect of poorly maintained spaces, Kodali et al. studied the implementation of urban park upgrades in impoverished New York City neighborhoods between 2017 and 2021^41^. They found a statistically significant increase in park utilization after public park renovations. Additionally, parents are often enthusiastic about physician-prescribed outdoor time, however, feel that access to green spaces is a barrier^42^. Further interventions to improve park quality and greenspace should be considered as a means to promote levels of physical activity in all children, including those particularly vulnerable to inactivity such as those with HCM.

The link between socioeconomic status and health outcomes is multi-faceted^43–47^, and we propose that the relationship between the environment and exercise capacity likely includes more than just the physical space in which these children live. For children with HCM, the decision to engage in exercise is complex and requires in-depth discussions with specialists, which patients from low COI neighborhoods may struggle to access. Furthermore, children with HCM require close supervision during exercise and sports participation, which is likely more feasible in resource-rich environments where multiple athletic trainers or teachers are present, and CPR training is more widespread^48,49^. Notably, there was a skew towards high or very high COI in our sample, despite most Philadelphia neighborhoods being categorized as low or very low COI. A map of various COI levels in Philadelphia and surrounding counties are shown in figure 3. Moreover, in our multivariate model, Black race was associated with higher exercise capacity, independent of poverty, urbanization or violent crime rates, despite Black patients typically having lower exercise capacity than non-Black patients^50–52^. We suspect that this may be indicative of referral bias, in that white children may be more likely to be referred for exercise testing than their Black counterparts of a comparable athletic ability. Taken together, these findings present a model wherein promoting healthcare access for children with HCM who grow up in low COI neighborhoods may be an important mechanism to increase physical activity and provide more equitable care.

**Figure 3.**
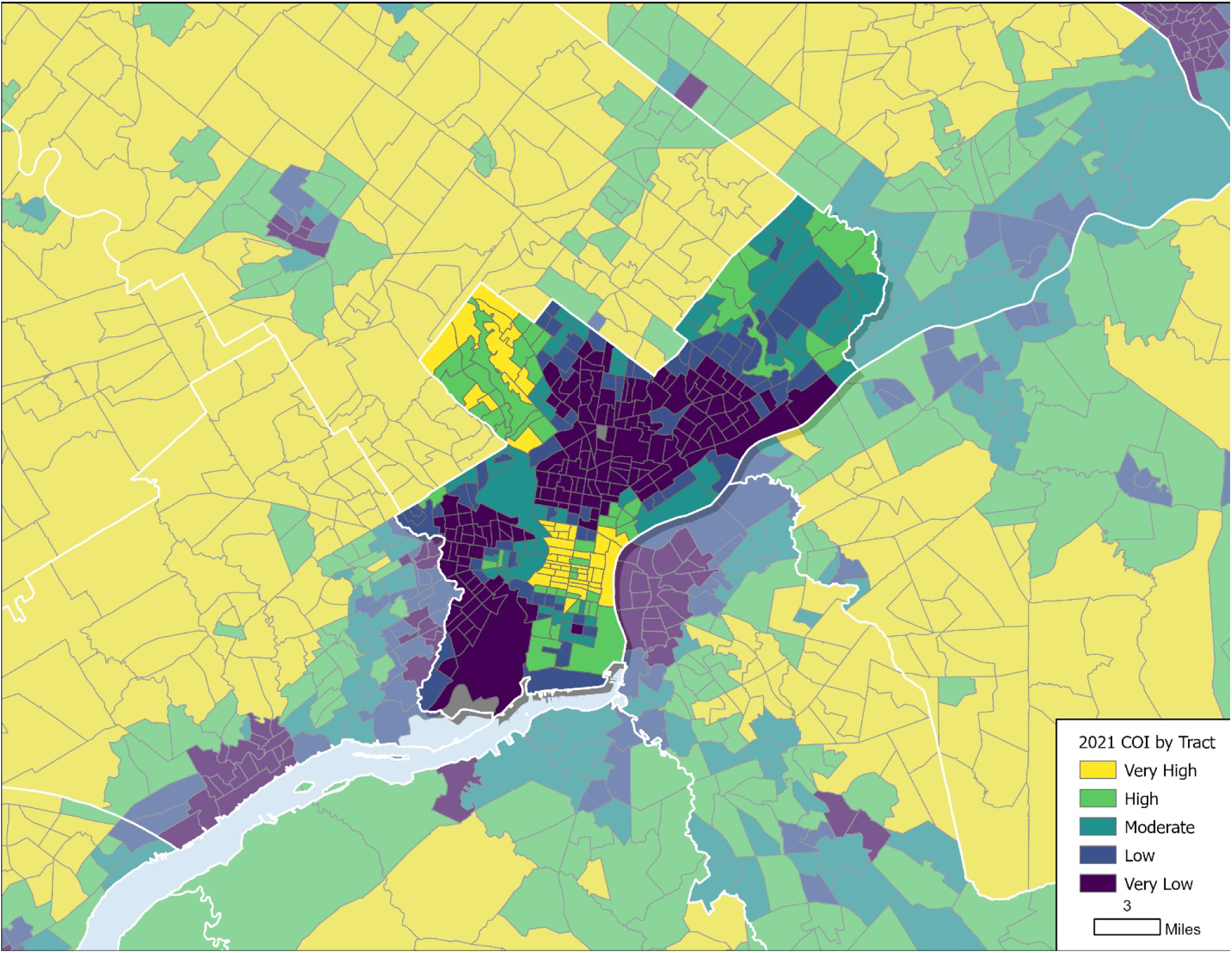
Map of Child Opportunity Index in Philadelphia and surrounding counties.

While many studies reveal that higher crime rates are associated with lower levels of physical activity^13,53–57^ and higher rates of obesity^58–60^, our study showed that higher crime rates were associated with better exercise capacity. The mechanism behind this relationship is not elucidated in this study, however, we suspect that this finding is potentially due to the limitations of the measure that we chose. Unlike COI, RUCA and other environmental exposures included in our analysis, crime rates were measured at the county level. Therefore, in Philadelphia County there is a single reported crime rate, despite the county’s highly heterogenous nature. Furthermore, the ability to assess the relationship between crime rates and exercise capacity may have been limited by our sample, which skewed towards high COI, therefore children from neighborhoods with higher crime rates may be underrepresented.

With the knowledge gained from this study, those who implement cardiac rehabilitation programs should consider childhood environment when designing exercise prescriptions. A child’s environment likely impacts both their exercise capacity and the ability to effectively implement exercise prescriptions. Providers should acknowledge patient financial resources and access to recreational spaces when implementing cardiac rehabilitation programs, as well as their own potential for referral bias. Use of virtual and home-based programs may offer a solution to overcome various neighborhood factors, particularly for those living in urban environments. Public health and public policies that improve the physical activity environment should be considered as interventions to improve cardiovascular health in children. Prospective studies that incorporate neighborhood-specific characteristics when designing interventions to measure the impact of cardiac rehabilitation on physical activity and exercise capacity are warranted to more conclusively define this interaction.

### Limitations

Our study is limited by its sample size and retrospective nature. The single-center study limits the generalizability of these findings. We created a conceptual model to represent environmental factors related to physical activity, however there is a complex interplay between environment, socioeconomic status, insurance status and patient geography that may not be entirely captured in our model. We attempted to utilize NDVI as an exposure, however this variable measures all vegetation and therefore does not directly translate to accessible green space such as parks. The accessible crime data was at a county level, as opposed to tract level, which limits the granularity of this exposure. Physical activity data was not available for this cohort therefore exercise capacity was used as a proxy of physical activity, and so the direct association of environment and physical activity may differ from what we found.

## Conclusion

This study utilizes a novel approach to understand the environmental determinants of exercise capacity in children with HCM. Those from lower COI, higher rates of urbanization and financially-poor neighborhoods had worse exercise capacity. We propose that neighborhood factors may impact levels of physical activity and play in these children, which in turn affects exercise capacity. Clinical implementation and studies of cardiac rehabilitation programs should account for patient resources and environment when designing and assessing the utility of exercise prescriptions.

## Data Availability

Data is available by request.

## Appendix

**Appendix Table 1.** Univariate regression analyses for adjusted peak VO2 (mL/kg/min)

| Exposures | Coefficient |
| --- | --- |
| COI – Education Z-score | 0.05 |
| COI – Health & Environment Z-score | 0.19 |
| COI – Social & Economic Z-score | 0.08 |
| Overall COI Z-score | 0.09 |
| COI - Education Score | 0.07 |
| COI – Health & Environment Score | 0.20 |
| COI – Social & Economic Score | 0.09 |
| Overall COI Score | 0.10 |
| National Walk Index | -0.01 |
| RUCA | 0.10 |
| Total Violent Crimes Rate | 0.09 |
| Murder Rate | 0.04 |
| Rape Rate | 0.12 |
| Robbery Rate | 0.09 |
| Aggravated Assault Rate | 0.08 |
| Total Property Crimes Rate | 0.03 |
| Burglary Rate | -0.04 |
| Larceny Rate | 0.03 |
| Motor Vehicle Theft Rate | 0.12 |
| Normalized Difference Vegetation Index | -0.21 |
| Index of Concentration at Extremes for Income | 0.05 |
| Index of Concentration at Extremes for Race | 0.17 |
| Index of Concentration at Extremes for Combined Income and Race | 0.12 |
| BMI | -0.24 |
| Age at Diagnosis of HCM | 0.09 |
| Peak septal thickness | -0.10 |
| Peak septal thickness Z-score | -0.11 |
| Peak LVOT gradient | -0.06 |
| Mean LVOT gradient | -0.08 |

None are statistically significant associations

**Appendix Table 2:**
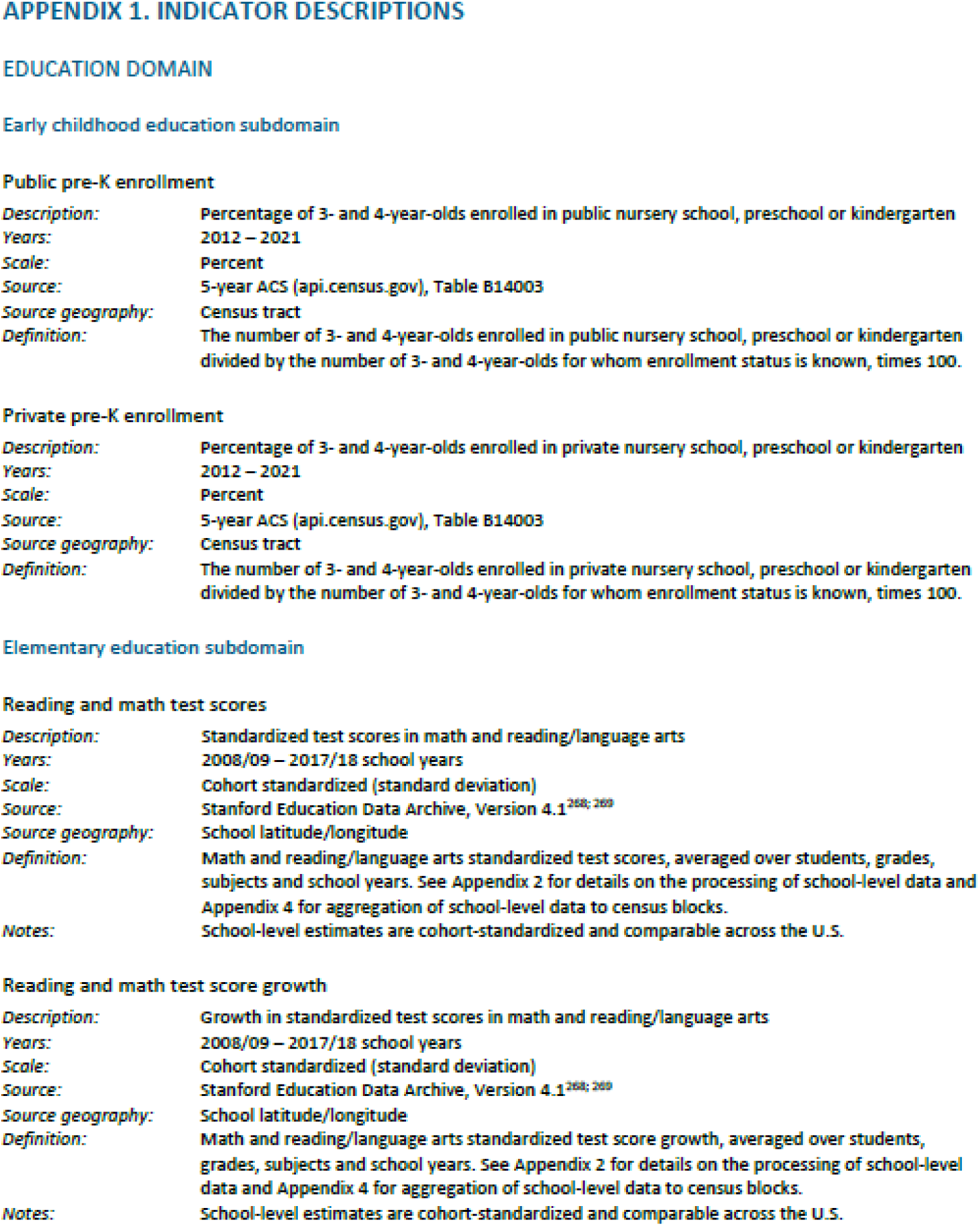
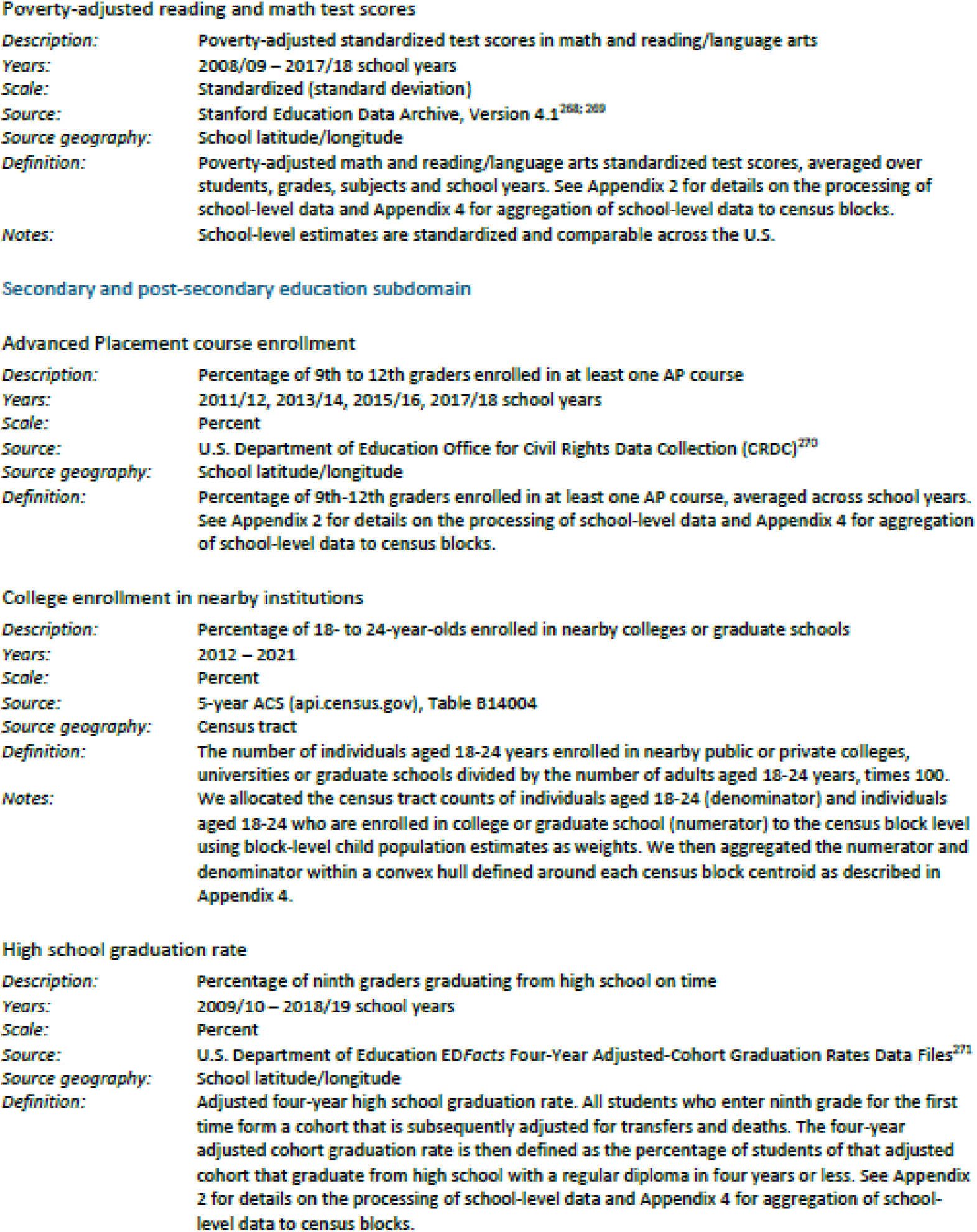
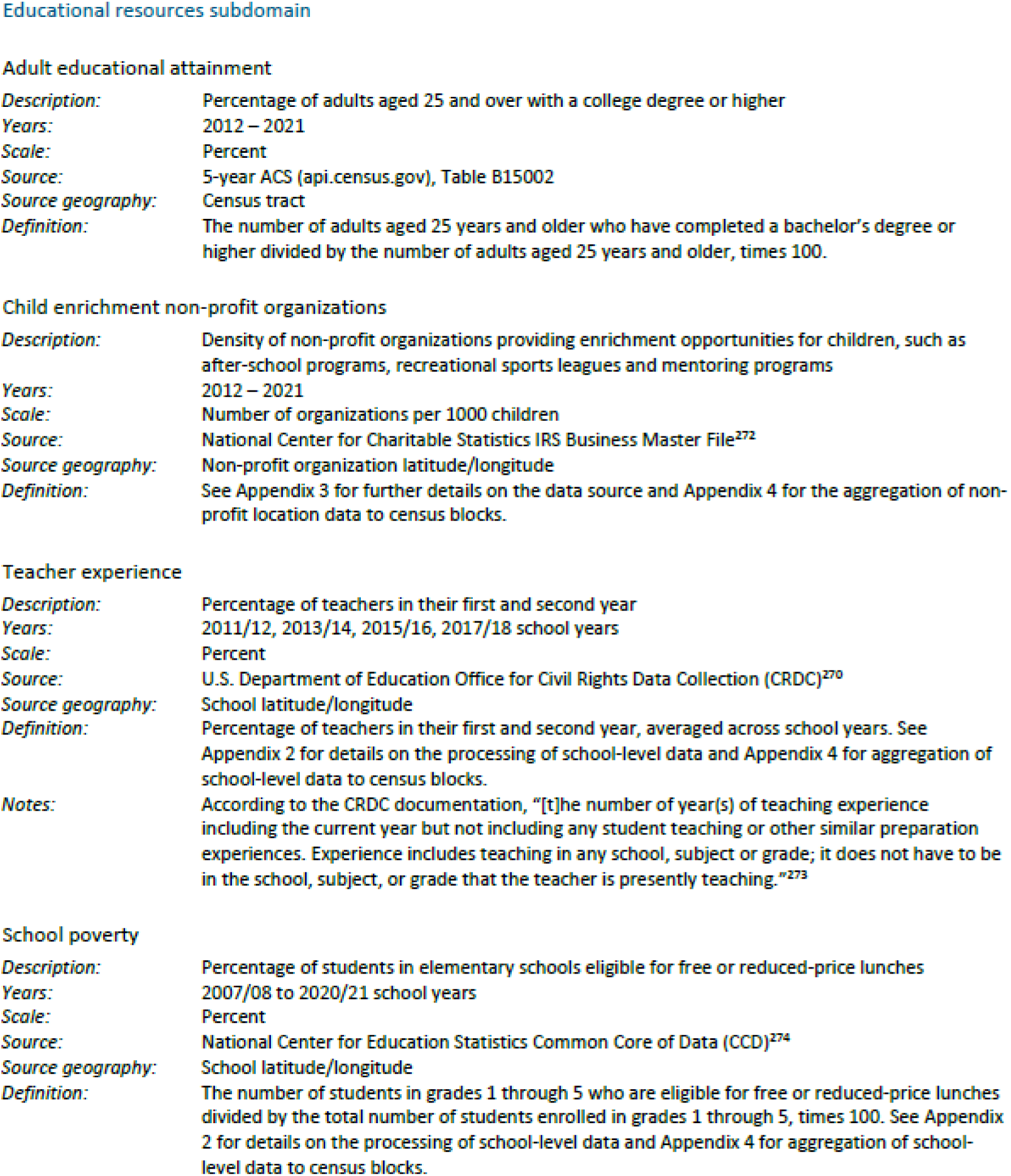
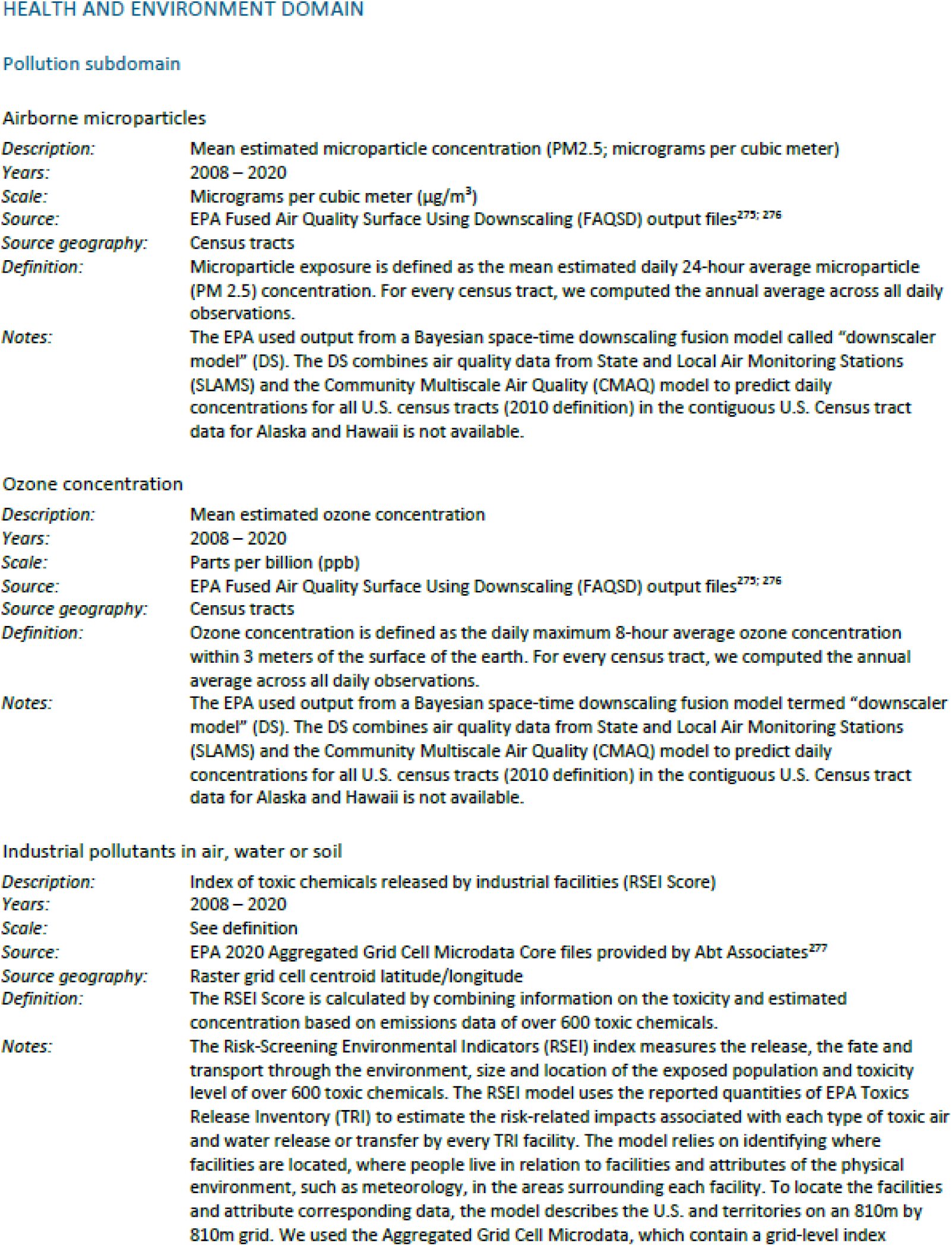
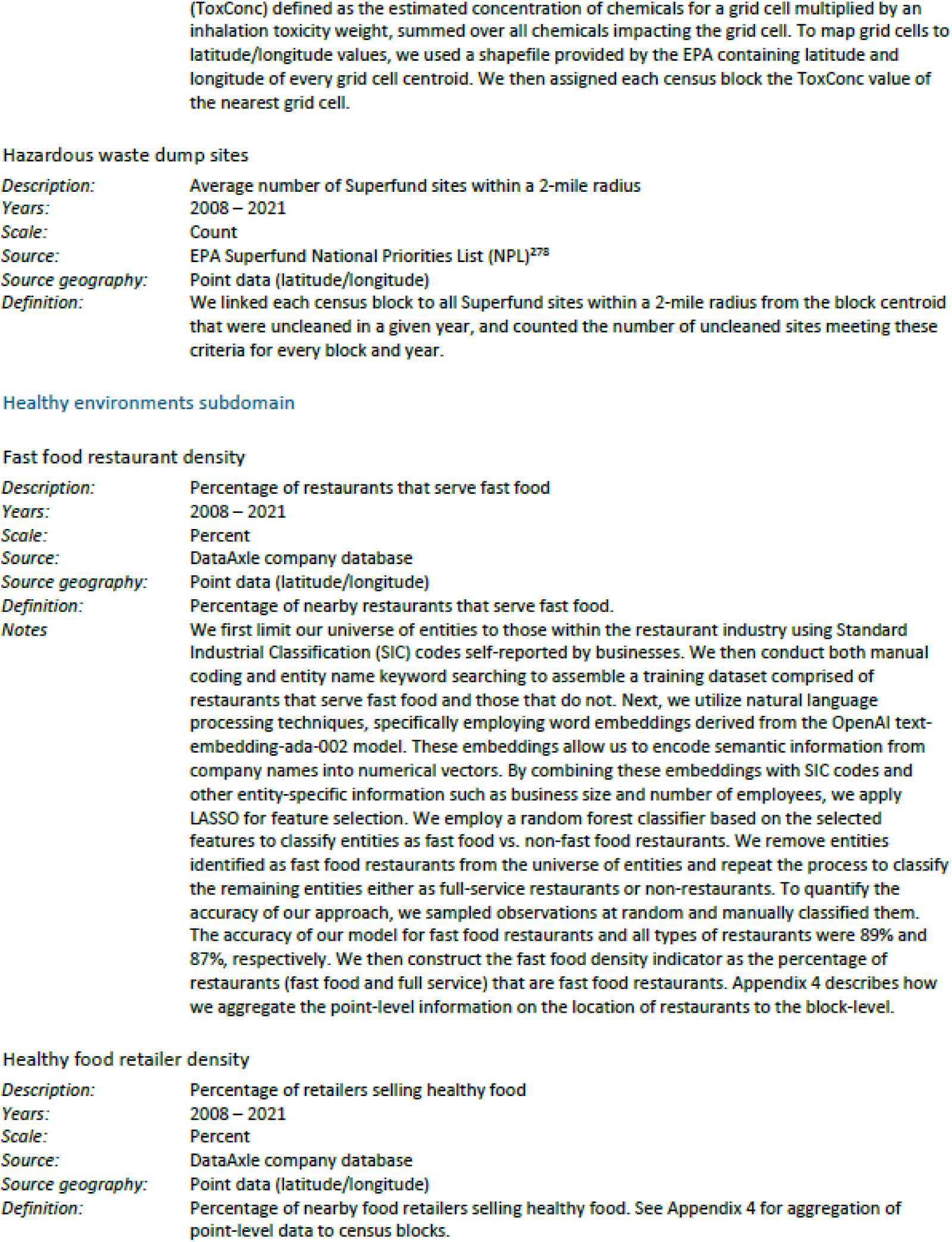
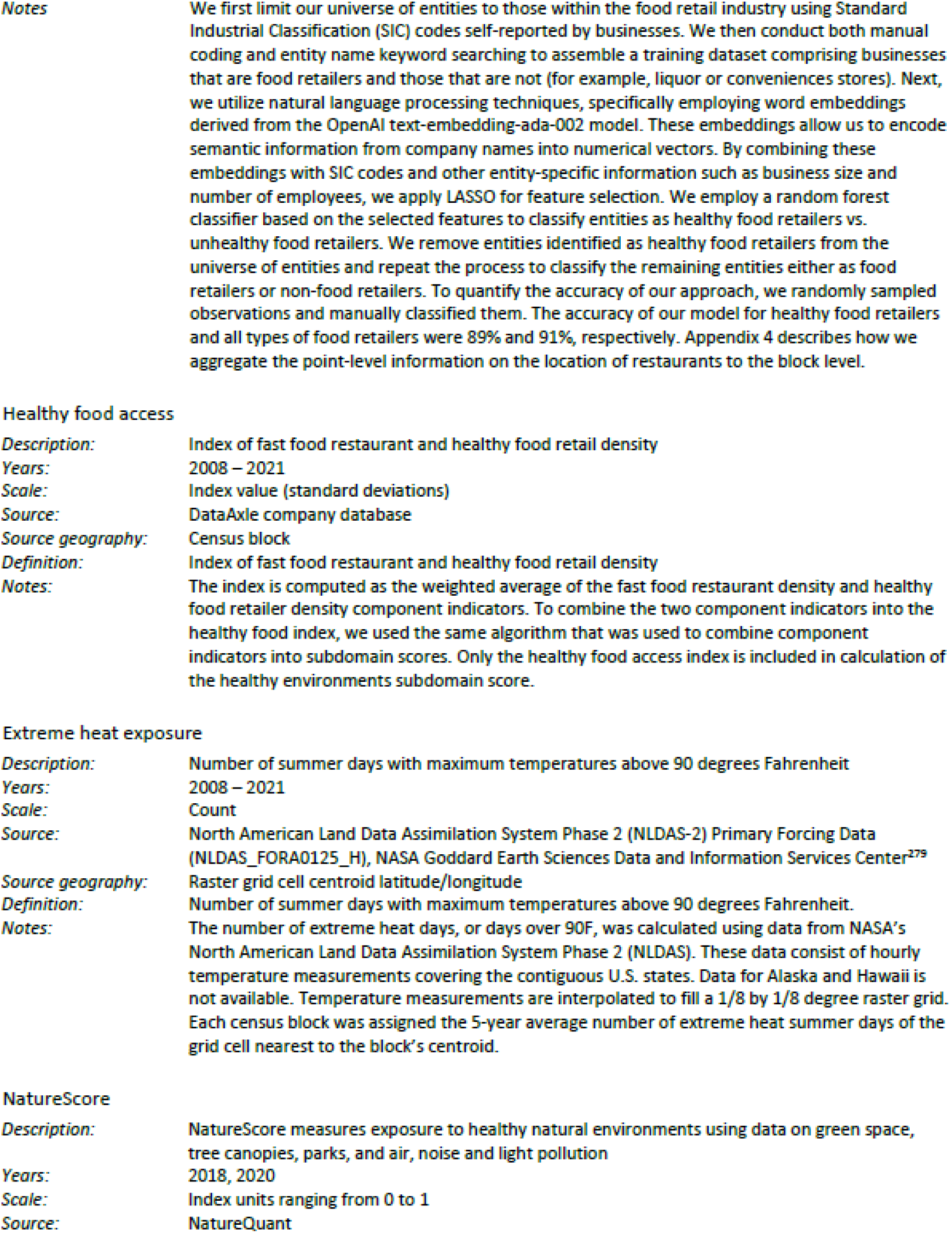
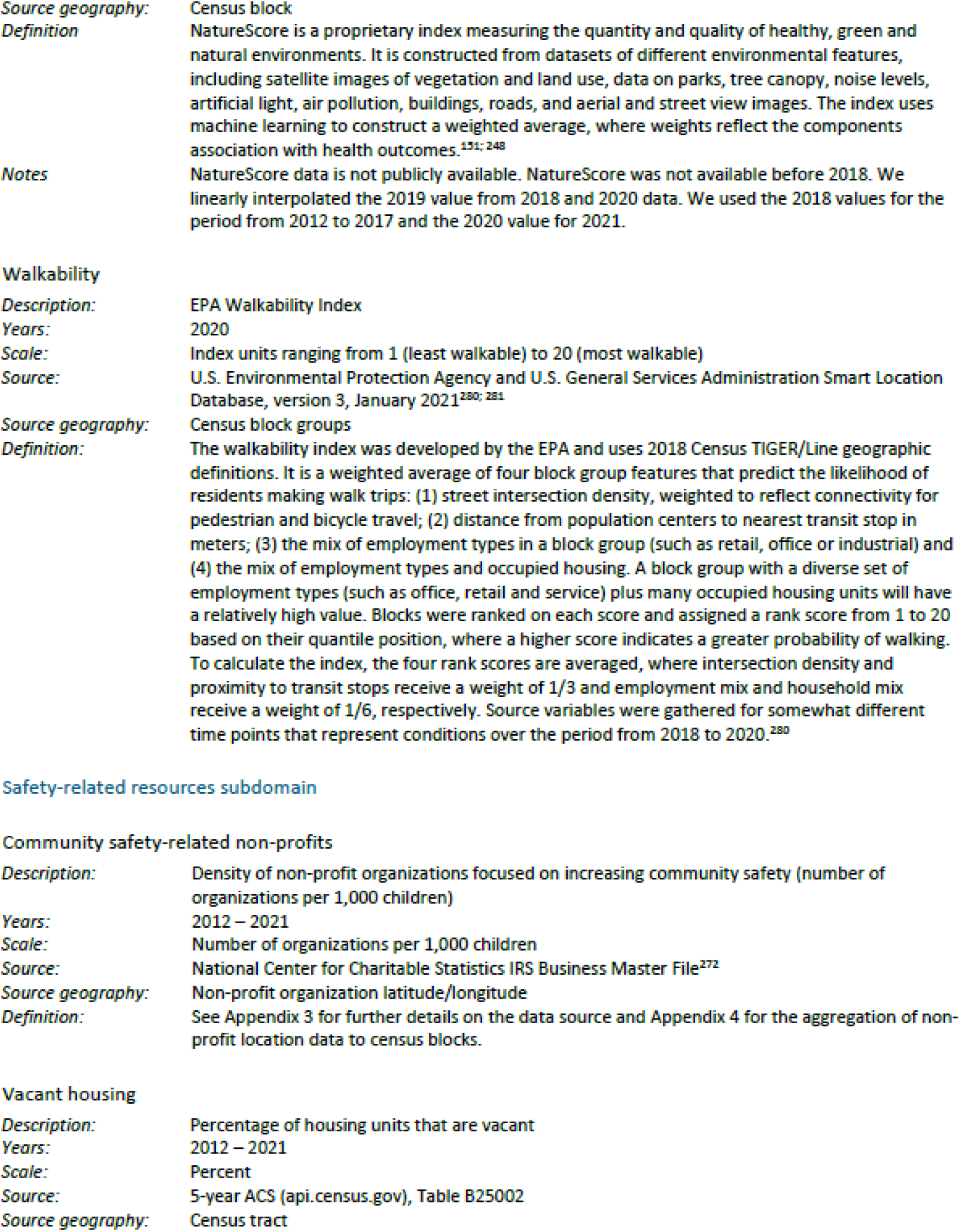
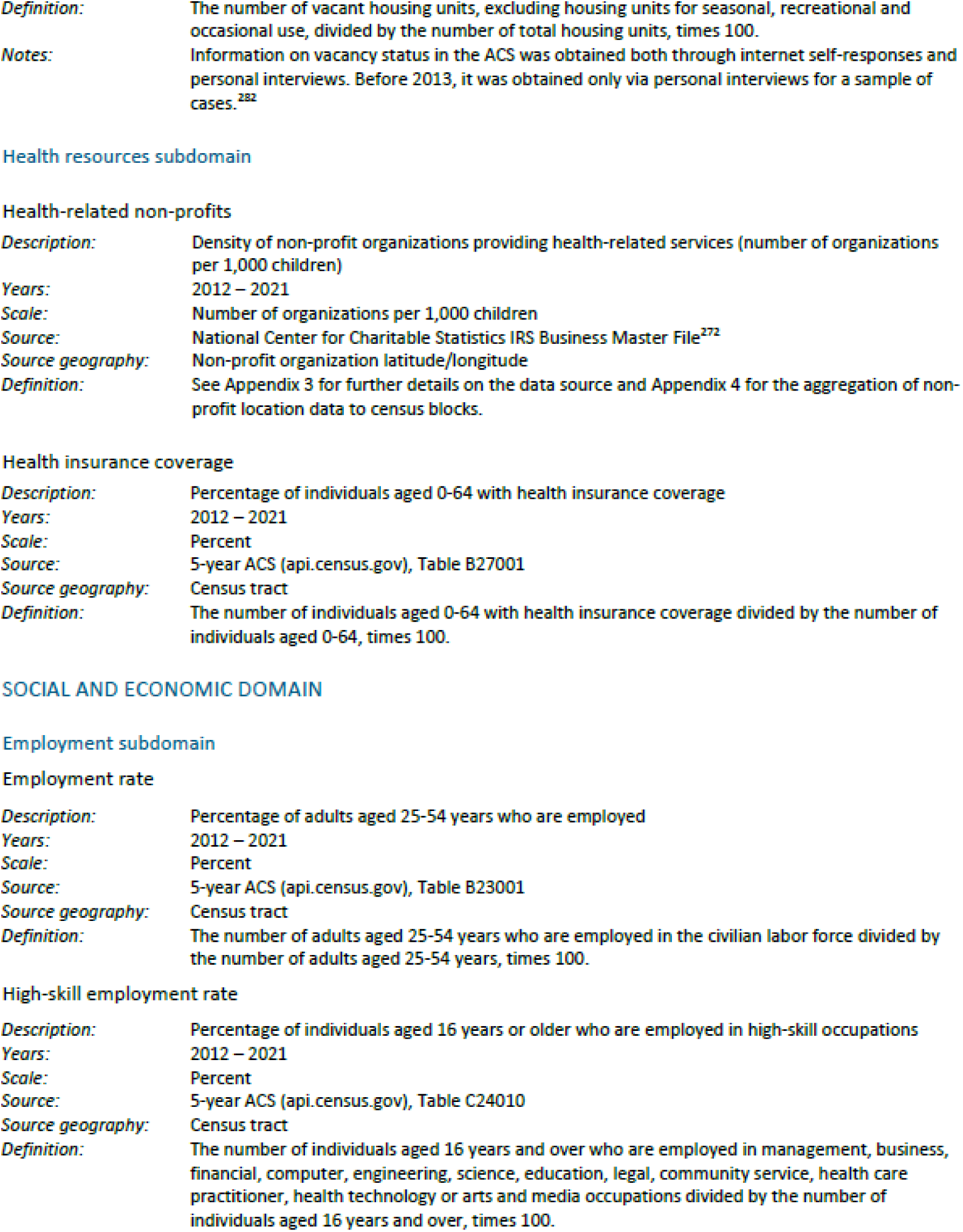
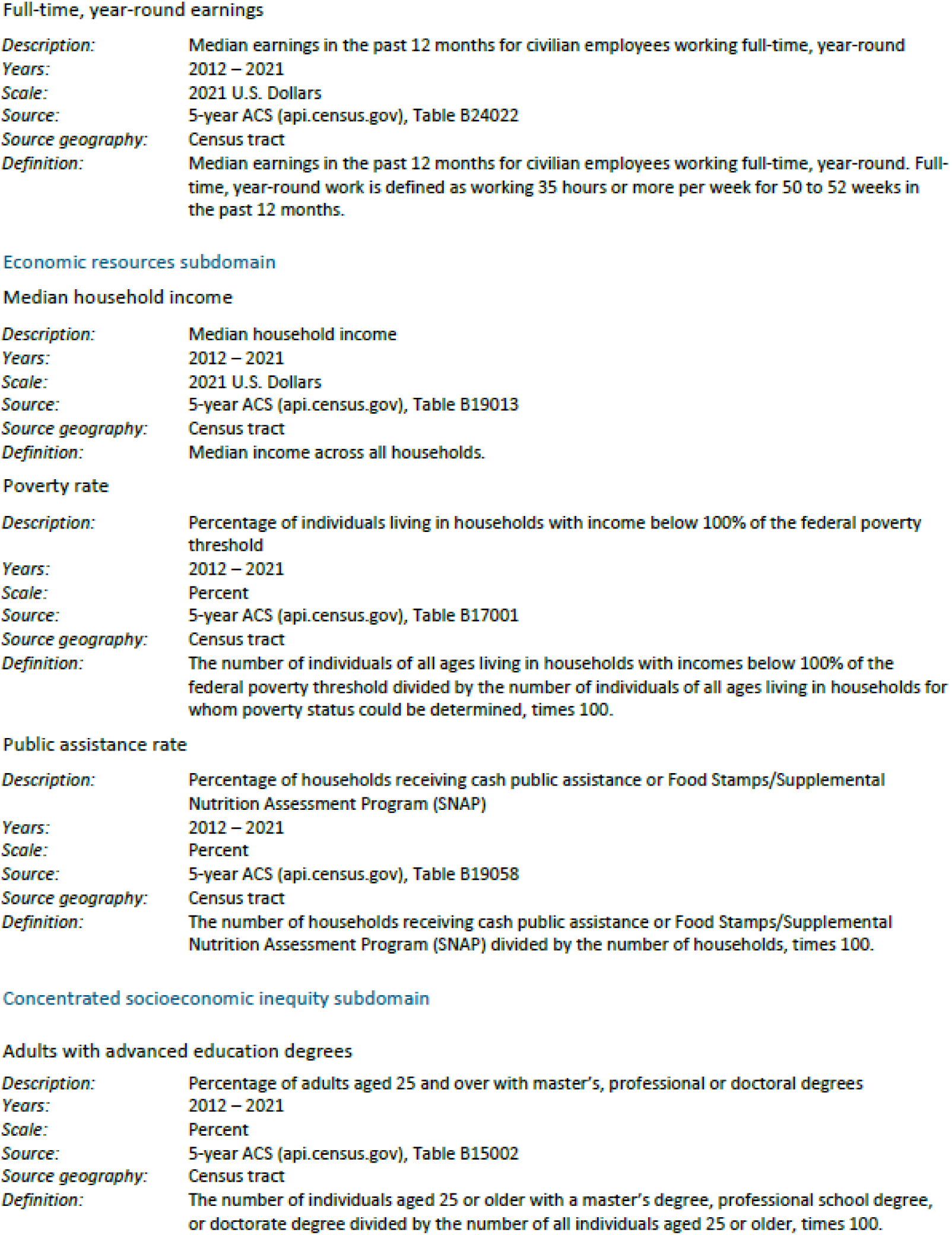
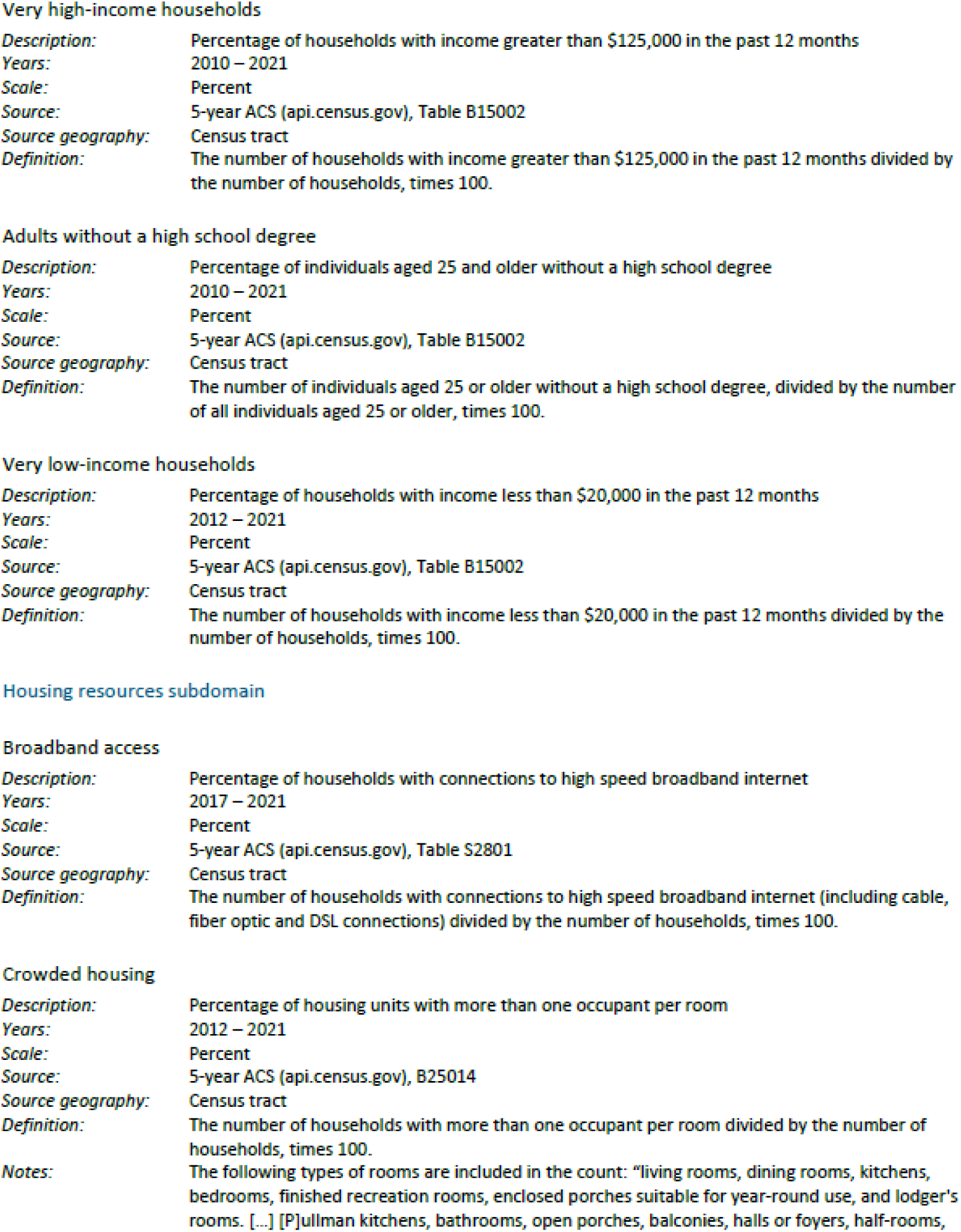
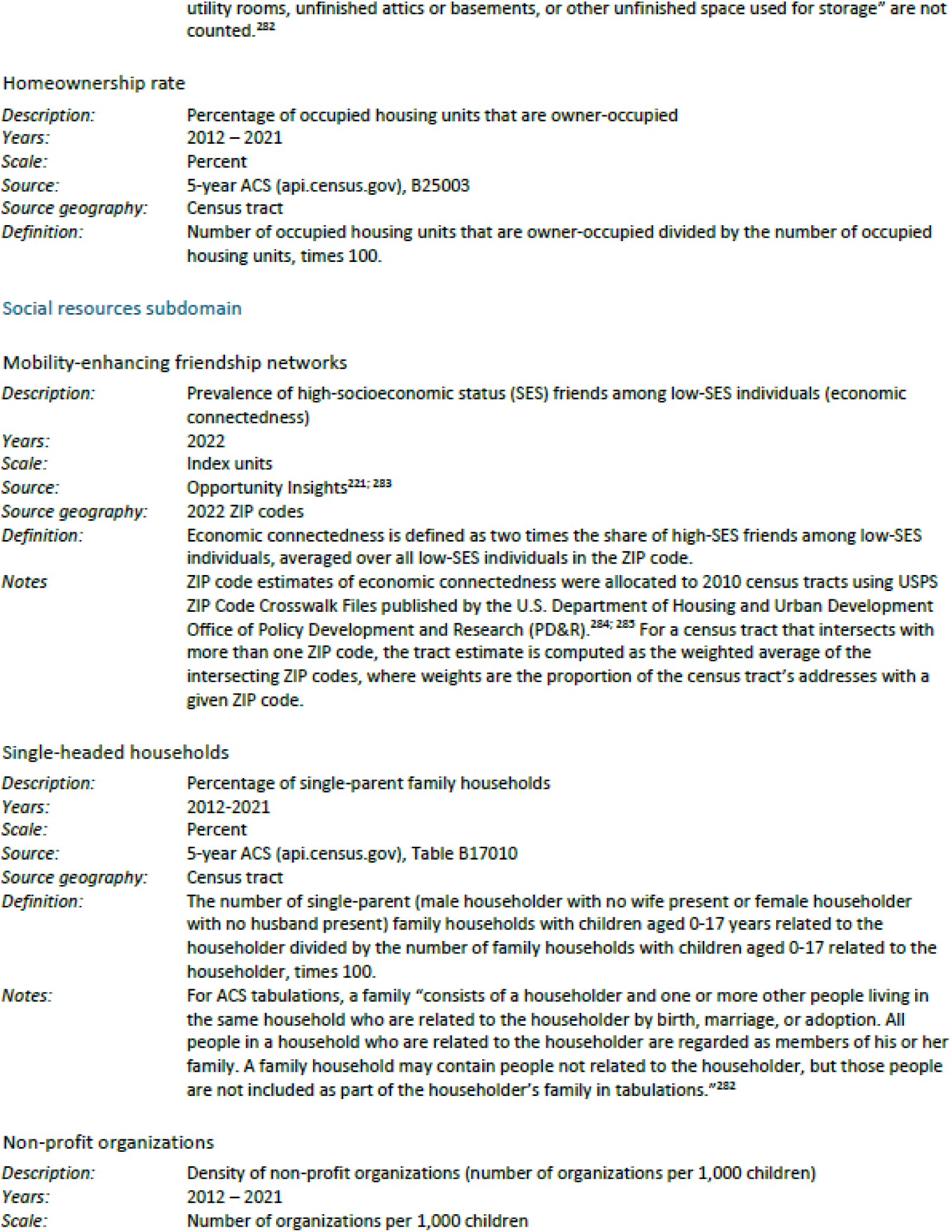
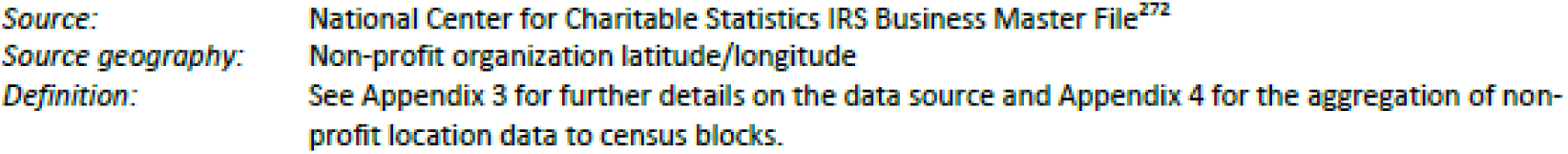
Child opportunity index subcategories.

